# Approaches for estimating COVID-19 vaccine effectiveness using observational data in administrative databases: a systematic review

**DOI:** 10.1101/2025.09.29.25336864

**Authors:** Jo Yi Chow, Zhi Jie Goh, Ruiqi Li, Darren Zi Yang Lim, Liang En Wee, David Chien Boon Lye, Kelvin Bryan Tan, Jue Tao Lim

## Abstract

**Background:** COVID-19 vaccine policy relied on observational vaccine-effectiveness (VE) studies conducted amid rapid variant turnover, evolving schedules, and shifting surveillance, yielding substantial heterogeneity in methodological approaches across studies. Prior reviews emphasised pooled or variant-specific VE, with limited attention to how methodological practice varied across countries and over time. Yet, understanding the landscape of methodological practices used during this period is essential for identifying opportunities to improve VE study design and conduct in future pandemic responses. This review systematically characterises the methodological practices in registry-based observational COVID-19 VE studies (2021–2024), documenting patterns in study design, statistical approaches, and analytical choices to establish an empirical foundation for methodological development in pandemic vaccine evaluation.

**Methods:** We ran a PRISMA-guided search of PubMed and Embase (via Ovid) from inception to Oct 14, 2024, for peer-reviewed observational studies estimating COVID-19 VE in routine (non-trial) settings that leveraged administrative/registry data (e.g., immunisation registries, laboratory/PCR databases, EHR/claims, hospitalisation/mortality registries, national-ID–linked datasets) and reported sufficient methodological detail to classify design, estimator, treatment of time, adjustment/matching/weighting, and sensitivity/validation checks. We excluded randomised trials; studies without administrative/registry data or confined to specialised populations; non-English publications; and duplicate analyses of the same cohort/time window. Descriptive summaries are presented overall, by calendar year, and by World Bank income group.

**Results:** 253 studies from 61 countries met eligibility; most were from high-income settings (187/253, 73.9%). The median publication lag was 257 days (IQR 157–421), lengthening from 141 days in 2021 to 673 in 2024, while median cohort size declined over time. Cohorts (46.6%) and test-negative designs (43.1%) dominated; target-trial emulations (2.0%) and quasi-experimental studies (1.2%) were uncommon. Logistic regression (56.1%) and Cox models (24.8%) comprised the majority of primary estimator. Adjustment emphasised demographics, comorbidity, calendar time, and geography; variables proximate to testing behaviour and exposure opportunity were less frequent. Most studies reported no matching/weighting (155/253, 61.2%); among those that did, exact matching predominated and weighting was rare. Sensitivity analyses were not described in 98/253 (38.7%) of studies. Endpoints concentrated on infection, hospitalisation, and mortality, while variant-resolved analyses waned as PCR testing and sequencing contracted.

**Conclusions:** Observational COVID-19 vaccine VE studies scaled rapidly where registries existed, but remained concentrated in high-income settings, relied on a narrow estimator set, and infrequently applied validity checks. Strengthening privacy-preserving linkages (including sequencing), aligning designs to target-trial principles with marginal weighting, and normalising a lean validity toolkit could enhance interpretability and policy relevance.

**Funding:** This research is supported by the National Research Foundation Singapore under its Clinician Scientist-Individual Research Grant (MOH-001572) and administered by the Singapore Ministry of Health’s National Medical Research Council. J.T.L. is supported by the Ministry of Education (MOE), Singapore Start-up Grant. L.E.W. is supported by the National Medical Research Council through the Clinician Scientist New Investigator Award.

## Introduction

Following identification of SARS-CoV-2 in late 2019, vaccines were developed at exceptional pace, with the first authorised for emergency use by December 2020 on the basis of randomised controlled trials (RCTs).(1,2) These trials, with high internal validity conferred by randomisation, established efficacy and safety sufficient for regulatory approval. The subsequent rapid deployment of COVID-19 vaccines, however, shifted the evidentiary burden from pre-licensure efficacy to post-authorisation vaccine effectiveness (VE), as RCTs could not feasibly encompass all age groups, dosing schedules, circulating variants, time-since-vaccination intervals, or clinically relevant outcomes.(3–7)

In routine settings, VE is an estimand defined by contrasts between specified vaccination strategies and comparator strategies over a stated risk period.(8) Its magnitude reflects both vaccine biology and context— population mixing, prior infection (including hybrid immunity), and the contemporaneous force of infection that varies over calendar time and place.(9) Administrative databases and national record linkages enabled VE estimation at scale,(10,11) enabling assessment of outcomes that were rare or underpowered in trials,(12) and facilitating analyses across demographic and clinical subgroups reflective of local epidemiology.(6,13) Such evidence guided policy decisions on vaccine allocation, booster timing, and the coordination of pharmaceutical and non-pharmaceutical interventions across successive pandemic phases.(14–16)

The context in which COVID-19 VE was estimated evolved rapidly. Successive variant waves altered baseline risk and immune escape; vaccine programmes progressed from primary series to multiple boosters and, later, bivalent formulations; and cumulative infections generated heterogeneous hybrid immunity.(17–19) In parallel, surveillance systems changed: testing intensity shifted, genomic sequencing capacity fluctuated, and the completeness and linkage of vaccination, testing, and clinical registries varied across jurisdictions.(20) These dynamics influenced eligibility to enter analysis, how exposures and outcomes are ascertained, which time scales were relevant, and the extent to which confounding, selection, and misclassification can be mitigated.

Therefore, a range of observational designs deployed during the pandemic, from case-based sampling frameworks (such as test-negative designs), prospective and retrospective cohorts, and quasi-experimental approaches. Although their data requirements and analytic implementations differ, these designs face shared challenges: confounding by indication and by prior infection;(21) calendar-time confounding during periods of rapidly changing incidence and variant emergence;(22) immortal-time and depletion-of-susceptibles bias;(23–25) differential testing and outcome misclassification;(26) and informative censoring.(27) Addressing these threats required explicit specification of estimands and comparator definitions, careful handling of calendar time and time-since-vaccination, and sufficient covariate information to support adjustment, matching, or weighting. The implementation of these approaches varies substantially across studies due to differences in population characteristics, data availability, and analytical choices, affecting the interpretability and comparability of published VE estimates.

Given these dynamics, a structured description of observational VE estimation across settings, calendar years, vaccines, comparators, and outcomes is needed to clarify the evidence base and its limits. Although several groups have synthesised VE magnitudes,(6,28,29) only one prior review focused on real-world study designs and was conducted mid-pandemic, before widespread booster programmes.(30) Accordingly, the present review provides a descriptive map of methodologies used in observational COVID-19 VE studies. We catalogue study settings, observation windows, vaccine and comparator definitions, study designs and estimators, covariate adjustment and matching/weighting, validation and sensitivity analyses, outcomes, and stated limitations. By organising common design and reporting elements, the review aims to support transparent interpretation of published VE estimates and to inform planning of future evaluations when capacity for randomised evidence is limited.

## Methods

### Literature search strategy

This systematic review mapped methodological approaches used in observational studies of COVID-19 vaccine effectiveness (VE); no meta-analysis of VE magnitudes was planned. Reporting followed PRISMA 2020 (**Supplementary Table 1a, 1b**). As the objective was methods characterisation rather than pooled effect estimation, a priori protocol registration (e.g., PROSPERO) was not undertaken.

We identified relevant studies through searches in PubMed and Embase (via Ovid) from database inception through October 14, 2024. The search strategy combined controlled vocabulary (MeSH/Emtree) and free-text terms aligned to the PICOS framework, with the search done for terms mentioned in the Title/Abstract (**Table 1**). Limits were applied to human, English-language, peer-reviewed publications; preprints and conference abstracts were excluded.

**Table 1.** Search strategy following the PICOS framework utilised for identifying peer-reviewed observational studies estimating COVID-19 vaccine effectiveness from administrative databases. Terms were categorised by patient/population (COVID-19 or related terms), intervention (vaccination and immunisation), comparator groups (various vaccination statuses), outcomes (measures of vaccine effectiveness), and observational study designs. Searches were restricted to human studies published in English, excluding preprints.

|  |  |
| --- | --- |
| **Patient/population** | “COVID-19” OR “SARS-CoV-2” OR “coronavirus” |
| **Intervention** | vaccin* OR immun* OR "booster" |
| **Comparison** | “unvaccinated” OR “unboosted” OR “partially vaccinated” OR "dose" OR "dosage" |
| **Outcome** | “vaccine effectiveness” OR “vaccine impact” OR “vaccine performance”<br>“retrospective study” OR “observational study” OR “cohort study” OR “case-control study” OR “test-negative design” OR “causal inference” OR “propensity score” OR “regression discontinuity” OR “instrumental variable” OR “difference-in-differences” |
| **Study design** | OR “survival analysis” |
| **Filter** | Humans, English, Exclude preprints |

Eligibility criteria required peer-reviewed studies that estimated COVID-19 VE in routine (non-trial) settings using observational designs that leveraged administrative or registry data (e.g., immunisation registries, laboratory/PCR databases, electronic health records or claims, hospitalisation and mortality registries, national ID–linked datasets) and reported sufficient methodological detail to classify design, estimator, adjustment, and sensitivity/validation checks. No restrictions were imposed on geography or outcomes. Randomised trials, immunogenicity- or laboratory-only studies, modelling/economic evaluations, case reports/series, studies without use of administrative/registry data, and duplicate analyses of the same cohort and window (retaining the most complete) were excluded. Additionally, to focus on broadly replicable approaches with typical administrative systems, studies confined to specialised subpopulations (e.g., healthcare workers, immunocompromised cohorts) were excluded, as were non-English publications due to limited accurate translation resources available to the team.

### Data extraction and analysis

Records were de-duplicated using unique identifiers (PMID/DOI) in EndNote. Two reviewers independently screened titles/abstracts against eligibility criteria, followed by full-text assessment for potentially eligible studies; discrepancies were resolved by consensus and, if needed, a third reviewer. A PRISMA flow diagram summarises identification, screening, eligibility, and inclusion (Figure 1). When multiple reports analysed the same underlying dataset but addressed distinct methodological questions (e.g., different designs, estimators, comparators, or endpoints), each report was retained as a separate methodological observation.

**Figure 1.**
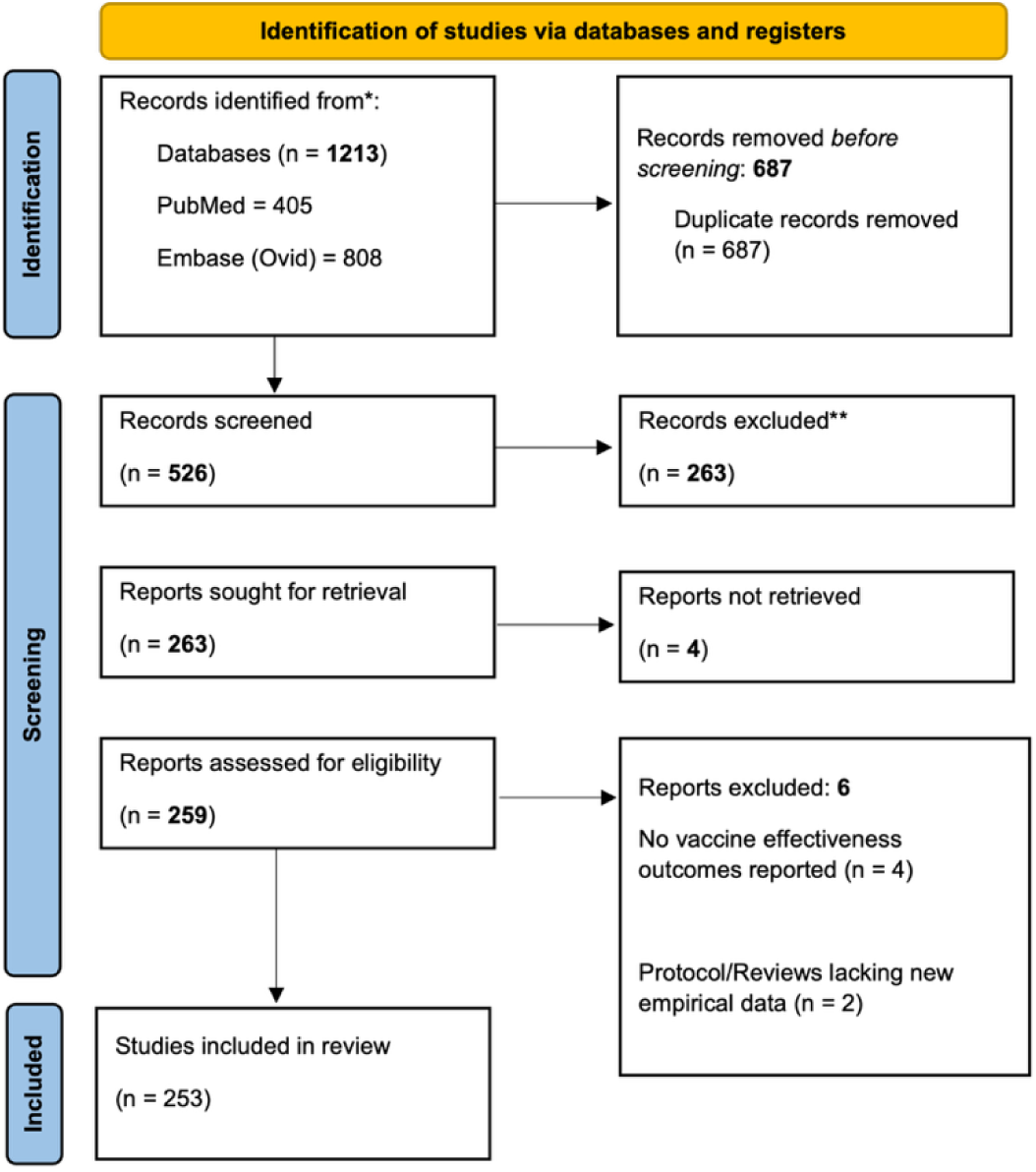
PRISMA reporting flowchart for studies on COVID-19 vaccine effectiveness

Four reviewers (JYC, ZJG, RL, DZYL) independently extracted data in duplicate using a standardised template refined through pilot testing on a subset of included studies (**Supplementary Table 2**). Briefly, extracted items included: study identifiers (publication year, country/region, observational period); study design with sample size/unit; exposure and comparator definitions; estimators/models; adjustment/matching/weighting variables and methods; outcomes and their ascertainment; sensitivity/validation checks; and author-stated limitations.

Operational definitions and decision rules were prespecified (**Supplementary Table 2**). Comparator contrasts were coded hierarchically as baseline (e.g., primary series vs unvaccinated; one dose vs unvaccinated), booster (e.g., boosted vs unvaccinated; boosted vs eligible-but-unboosted), and platform/variant comparisons; studies could contribute multiple contrasts. Publication lag was defined as days from the start of the observational period to the journal publication date. Study setting income groups followed the 2023 World Bank Atlas method.(31)

A hierarchical coding framework supported consistent cataloguing (**Supplementary Table 2**). Sensitivity/validation analyses were grouped into nine domains: alternate exposure definitions; alternate outcome definitions/ascertainment; alternative model specifications; comparator re-definitions; calendar-period restrictions; negative controls; eligibility-criteria changes; missing-data handling; and checks for unmeasured confounding (including E-values). Reported covariates were mapped into ten themes: demographic; temporal; clinical comorbidities/health status; geography/residence; socioeconomic; prior SARS-CoV-2 infection and vaccination history; healthcare utilisation; exposure risk/occupation; testing behaviour/prior infections; and household/living conditions.

As the objective was to characterise methods rather than pool VE, formal instrument-based risk-of-bias appraisal (e.g., ROBINS-I) was not applied. Instead, presence or absence of assumption checks, together with author-stated limitations, was catalogued as an indicator of methodological rigour. Descriptive summaries were produced overall, by calendar year (2021–2024), and by World Bank income group.(31) Categorical variables are presented as counts (percentages) and continuous variables as median (in publication lag) or totals (with cohort size) and corresponding interquartile ranges (IQR). Analyses and visualisations were generated in R (version 2023.06.1+524). The full list of studies and corresponding characteristics is available as **Supplementary Table 3**.

## Results

We identified 1,213 articles through the database search, with 405 identified from PubMed and 808 from Ovid, as documented in **Figure 1**. No additional articles were identified from searching reference lists. After removal of duplicates (687) and exclusion of studies based on screening the abstract (263), full text (6), or non-retrievability (4), a total of 253 eligible studies were identified. Of the 6 studies excluded during full text screening, 4 did not evaluate vaccine effectiveness,(32–35) and 2 were protocols or reviews lacking new empirical data.(36,37)

Figure 2 illustrates the global distribution of the 253 observational COVID-19 VE studies across 61 countries (grey denotes no identified studies). Regionally, Asia accounted for the greatest share (n = 87),(38–118) followed by Europe(119–183) and North America(184–250) (n = 67 each), South America (n = 18),(251–267) Africa (n = 6)(268–273) and Oceania (n = 5);(274–278) three studies spanned more than one continent (Europe & South America, n = 3).(279–281) Study counts were highest in the United States (n = 47),(185,186,188–195,197–200,202,204–206,208,210–213,215–217,219–221,224–226,229,231–233,235,237–243,247–249) the United Kingdom (n = 23)(131,133,135,138,140,155,157,158,164,170,172,179) and Mainland China (n = 17).(45,50,51,58,65–67,70–72,94,95,102,105,113,118,282) In contrast, Central Asia, much of sub-Saharan Africa and large areas of South America remain largely unrepresented in the observational VE evidence base.

**Figure 2.**
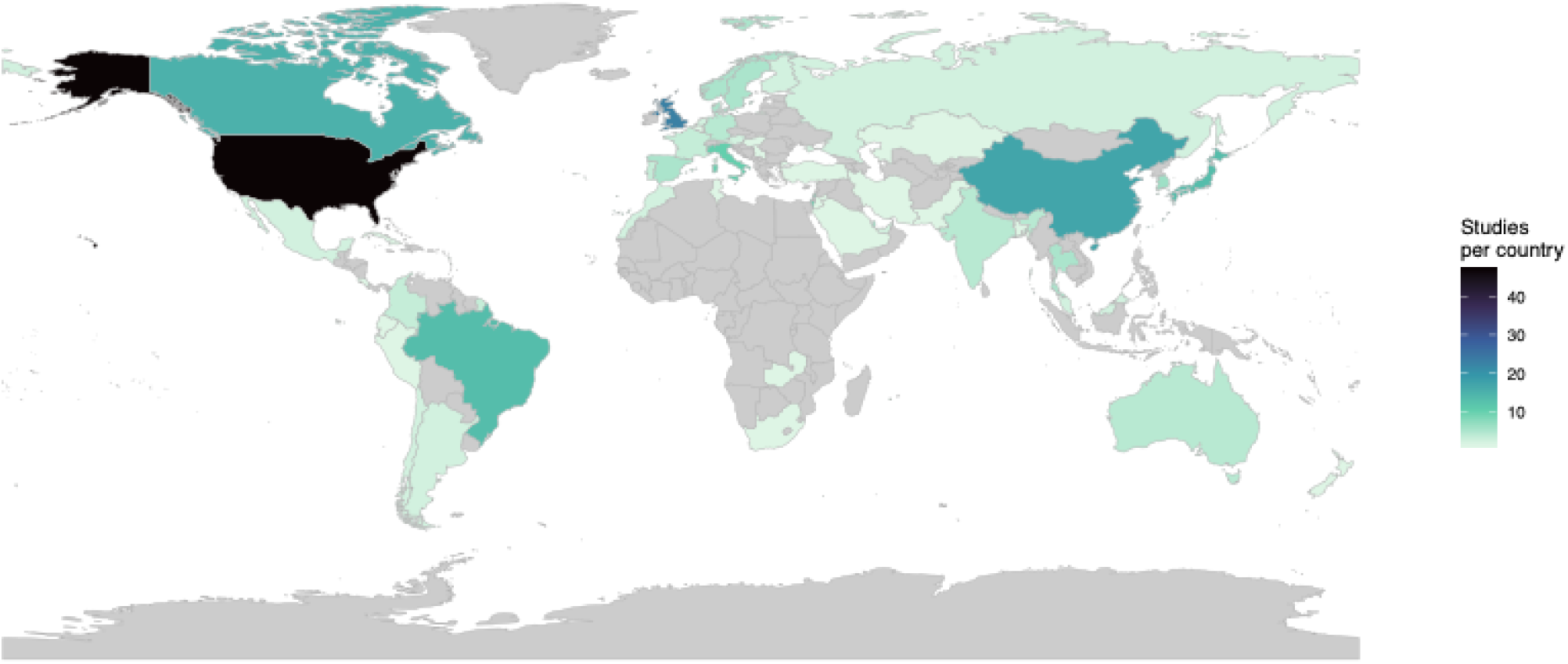
Global distribution of COVID-19 vaccine effectiveness observational studies (n = 253). Countries are shaded according to the number of included studies (darker colours = more studies; grey = no studies identified).

**Table 2** summarises the methodological and epidemiological features of the 253 included studies. Publications spanned 2021–mid-2024, with a median publication lag of 257 days (IQR 157–421). Most were conducted in high-income settings (187/253, 73.9%), peaked in 2022 (113/253, 44.7%), and were predominantly retrospective (219/253, 86.6%). Cohort (118/253, 46.6%) and test-negative case-control designs (109/253, 43.1%) dominated; quasi-experimental designs were rare (3/253, 1.2%). For estimators, logistic regression (147, 56.1%)and Cox proportional hazards models (65, 24.8%) together comprised ∼81% of uses. Overall, the 253 studies collectively evaluated 364 334 082 (IQR 9 813–960 307) individuals, 194 220 (IQR 58 880–135 340) households, and 117 951 198 (IQR 38 995–444 220) COVID-19 RAT/PCR tests.

**Table 2.**
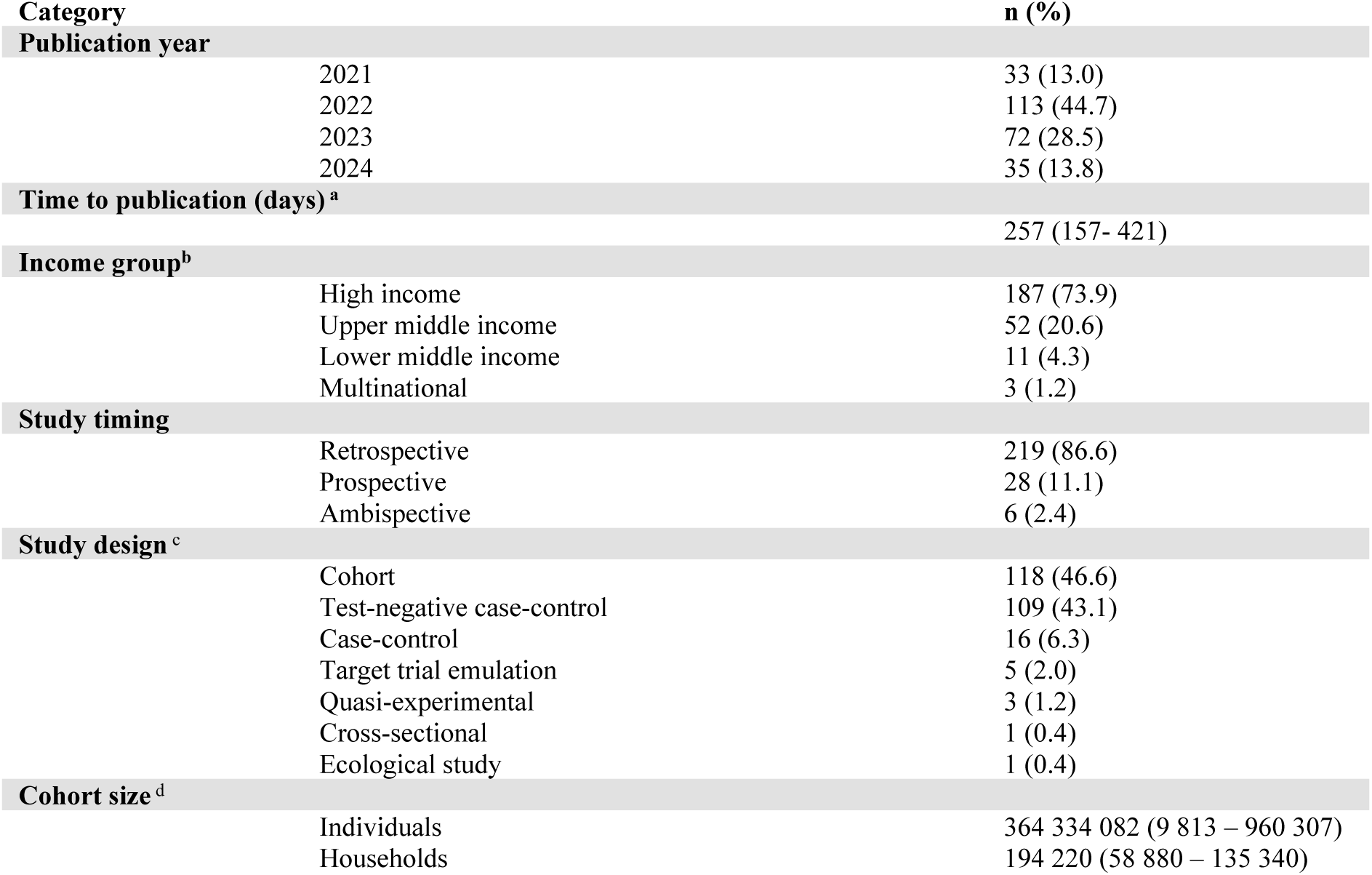
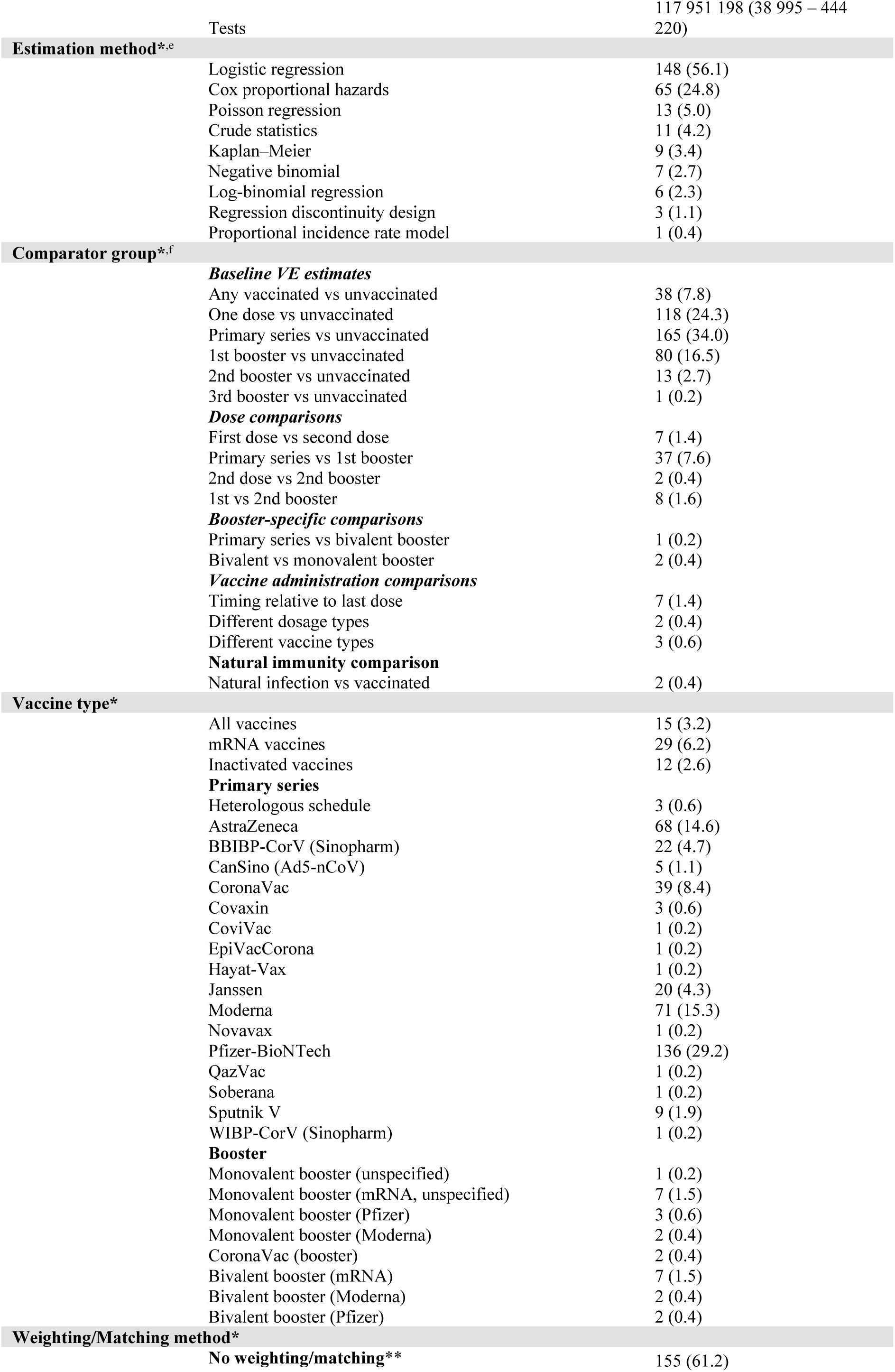
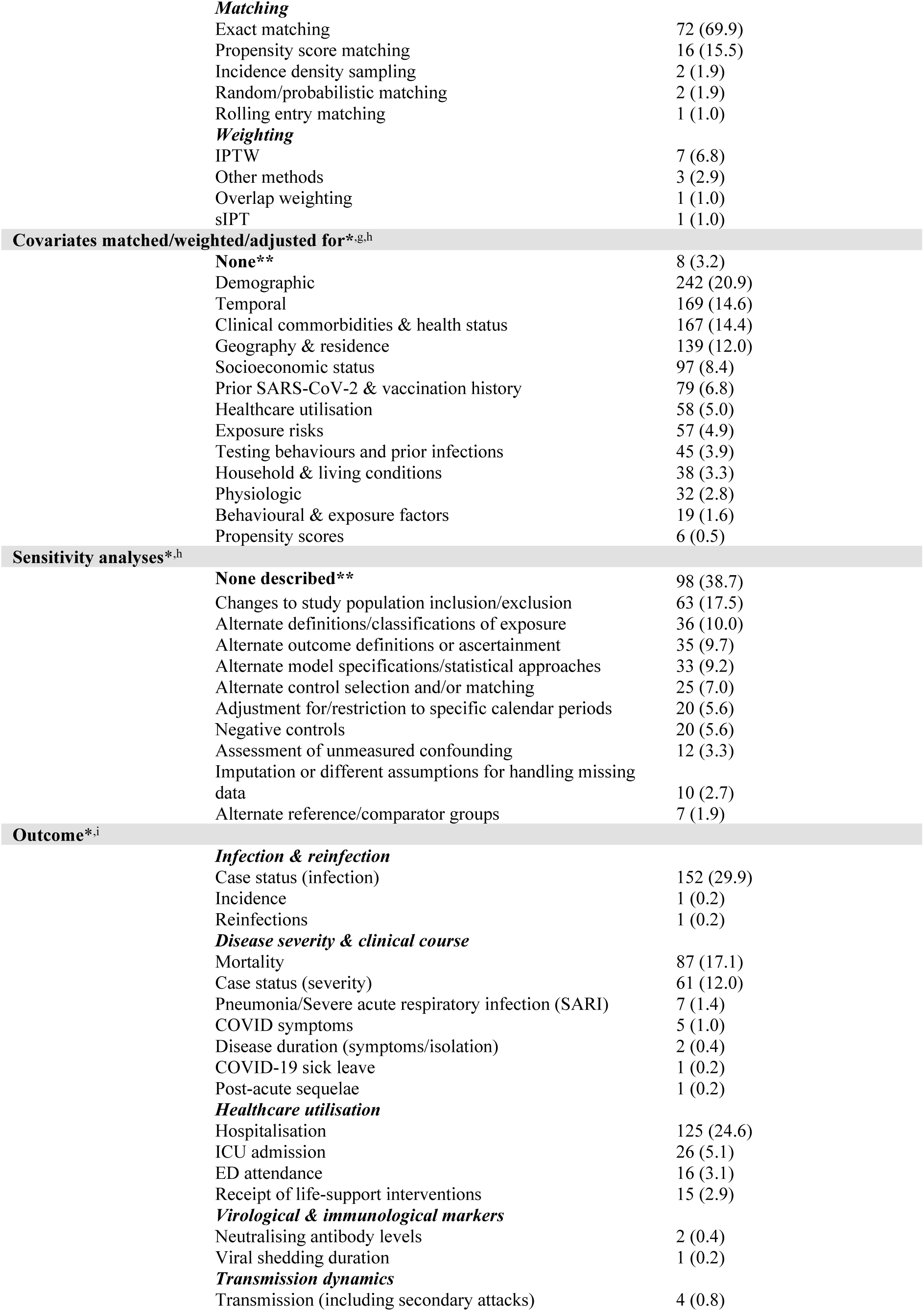
Overall characteristics of the included studies (N = 253). Values are n (%) unless stated; medians and totals are shown with interquartile ranges (IQR). Abbreviations: VE, vaccine effectiveness; IQR, interquartile range; ED, emergency department; ICU, intensive care unit; SARI, severe acute respiratory infection; IPTW, inverse probability of treatment weighting; sIPT, stabilised inverse probability of treatment weights. See Supplementary Table S2 (codebook) for the full mapping from study-reported variables to covariate domains and sensitivity-analysis categories. * Percentages are calculated within each domain. For single-select domains (e.g., publication year, study timing, study design), the denominator is the number of studies contributing data to that domain (typically N=253). For multi-select domains marked with an asterisk (*) (i.e. estimation method, comparator group, vaccine type, weighting/matching, covariates, sensitivity analyses, outcomes), the denominator is the total number of domain-specific observations after expanding multi-category contributions (e.g., distinct contrasts, vaccine products, covariate domains). Thus a study may contribute >1 observation within a domain; totals may exceed 100% across subcategories, and minor deviations from 100% reflect rounding. ^a^ Time to publication is calculated as days from the start of study follow-up to its journal publication date, reported as median (IQR). ^b^ Income group per World Bank country income classifications 2023; (31) ‘Multinational’ indicates studies spanning countries in more than one income group. ^c^ Assigned from authors’ primary analysis; ‘test-negative’ treated as a case-control variant; ‘target trial emulation’ used only when explicitly stated. If a study reported multiple designs, classification follows the analysis prespecified or labelled ‘primary’ by the authors. ^d^ Cohort size is reported as the sum total with across studies with its corresponding IQR by unit of analysis (individuals, households, tests). Totals and IQRs computed only among studies reporting that unit. ‘Tests’ counts specimens/episodes, not unique individuals. ^e^ Method reflects the primary effect estimator; ‘logistic regression’ includes conditional and mixed-effects; “crude statistics” includes unadjusted risk/odds/rate comparisons; ‘Kaplan–Meier’ denotes unadjusted survival contrasts. ^f^ ‘Primary series’ follows country-appropriate schedule; ‘bivalent’ = Omicron-containing products; ‘any vaccinated’ = ≥1 dose with study-specific risk window (typically ≥14 days). ^g^ ‘Propensity score’ under covariates means entered as a covariate, not used for matching/weighting. ^h^ Percentages are computed within domain [sensitivity analyses (n=291), weighting/matching (n=103), covariates (n=1,148)] except rows marked **, i.e. ‘No weighting/matching’, ‘None’ (covariates), and ‘None described’ (sensitivity analyses/validation). Marked rows use the total number of studies (N=253), i.e., the share of studies with no such feature. Each study can only contribute to a particular category once. ^i^ Definitions follow each study; ‘infection’ = laboratory-confirmed (PCR or antigen) unless stated; ‘hospitalisation’ and ‘ICU’ as reported (COVID-related vs all-cause may vary); ICU includes HDU where combined; severity definitions follow each study’s criteria (WHO/ICD or country-specific). See Supplementary Table S2 (codebook) for the full mapping from study-reported sensitivity analyses to outcome categories.

Comparator structures most often contrasted primary series vs unvaccinated (165, 34.0%) or one dose vs unvaccinated (118, 24.3%); contrasts involving booster doses were less common (e.g., first booster vs unvaccinated: 80, 16.5%; primary series vs first booster 37, 7.6%), as were natural-immunity comparisons (2, 0.4%). Vaccine products most frequently evaluated were Pfizer-BioNTech (136, 29.2%), Moderna (71, 15.3%), and AstraZeneca (68, 14.6%), with CoronaVac (39, 8.4%) and BBIBP-CorV (22, 4.7%) also represented.

Most studies used no matching or weighting (155/253, 61.2%). Among analyses that did, exact matching predominated (69.9%), followed by propensity-score matching (15.5%); weighting approaches included IPTW (6.8%), overlap weighting (1.0%), and stabilised IPT weights (1.0%). Covariate control most often covered demographic (242, 20.9%), temporal (169, 14.6%), and clinical comorbidity/health-status domains (167, 14.4%).

About three in five studies reported at least one sensitivity analysis (i.e., 61.3% had ≥1; 98/253, 38.7% reported none). The most frequent checks involved changes to inclusion/exclusion criteria (63, 17.5%), alternative exposure definitions (36, 10.0%), alternative outcome definitions/ascertainment (35, 9.7%), and alternative model specifications (33, 9.2%). Outcomes (multi-select) most commonly included infection (152, 29.9%), hospitalisation (125, 24.6%), and mortality (87, 17.1%), with fewer analyses of ICU admission (26, 5.1%), ED attendance (16, 3.1%), and immunological markers (≤0.4% each). Cohort size medians by unit (individuals, households, tests) are reported in Table 2. Commonly cited limitations included residual confounding (e.g., incomplete data on prior infection or healthcare-seeking), exposure/outcome misclassification, and variability in geographic coverage, variant surveillance, and follow-up windows.

Stratification by income group revealed marked differences, particularly in publication speed and sample scale (**Table 3**). High-income country (HIC) studies were published fastest among country strata with a median of 245 (IQR 144–391) days, followed by lower-middle-income (LMIC) studies at 338 (IQR 173–398) days and upper-middle-income (UMIC) studies with 391 (IQR 200–598) days; multinational analyses had the shortest lag overall with a median of 111 (IQR 90–187) days. Median cohort sizes likewise tracked resources: HIC studies enrolled a median of 178 153 individuals (IQR 17 898–1 096 216) versus 16 201 (IQR 3 353–278 642) in UMIC and 1 731 (IQR 1 315–4 414) in LMIC; household-unit cohorts were observed only in HIC (median 97 110 [IQR 58 881–135 340]), and studies using tests were largest in HIC (median 143 112 [IQR 47 951–444 220]), with UMIC far smaller (median 10 077 [IQR 9 018–104 018]).

**Table 3.** Characteristics of included studies by World Bank income group. Values are n (%) within each income stratum unless stated; medians are shown with interquartile ranges (IQR). Column denominators are the number of studies in each stratum: High-income N = 187; Upper-middle N = 52; Lower-middle N = 11; Multinational N = 3. Definitions for study design categories, estimation methods, comparator mappings, vaccine product labels, and outcome groupings follow those in Table 2. Abbreviations: VE, vaccine effectiveness; IQR, interquartile range; ED, emergency department; ICU, intensive care unit; SARI, severe acute respiratory infection; IPTW, inverse probability of treatment weighting.; sIPT, stabilised inverse probability of treatment weights. * denotes multi-select domains. These use the domain-specific observation total within each income group stratum as the denominator; studies may contribute >1 observation; minor deviations reflect rounding. ^†^Denominator rule. For the rows “No weighting/matching,” “None” (covariates), and “None described” (sensitivity analyses), percentages use the study counts by income group (HIC N=187; UMIC N=52; LMIC N=11; Multinational N=3). All other rows are calculated within their section, excluding those “None/No …” rows, using these denominators: **Matching/Weighting**—HIC n=83, UMIC n=17, LMIC n=4, Multinational n=1; **Covariates**—HIC n=906, UMIC n=150, LMIC n=34, Multinational n=63; **Sensitivity analyses**—HIC n=210, UMIC n=42, LMIC n=3, Multinational n=6. (N = number of studies; n = within-section total.)

| Category | Income group |  |  |  |
| --- | --- | --- | --- | --- |
|  | High income<br>(N = 187) | Upper middle income<br>(N = 52) | Lower middle income<br>(N = 11) | Multinational<br>(N = 3) |
| Publication year |  |  |  |  |
| 2021 | 28 (84.8) | 3 (9.1) | 2 (6.1) | 0 (0) |
| 2022 | 84 (74.3) | 19 (16.8) | 8 (7.1) | 2 (1.8) |
| 2023 | 52 (72.2) | 19 (26.4) | 0 (0) | 1 (1.4) |
| 2024 | 23 (65.7) | 11 (31.4) | 1 (2.9) | 0 (0) |
| Median time to publication |  |  |  |  |
|  | 245 (144–391) | 391 (200–598) | 338 (173–398) | 111 (90–187) |
| Median cohort size (median [IQR]) |  |  |  |  |
| Individuals | 178153 (17898–1096216) | 16201 (3353–278642) | 1731 (1315–4414) | 25181752 (15506981–34856522) |
| Households | 97110 (58881–135340) | NA | NA | NA |
| Tests | 143112 (47951–444220) | 10077 (9018–104018) | NA | 630944 (630944–630944) |
| Study type |  |  |  |  |
| Cohort | 95 (50.8) | 20 (38.5) | 2 (18.2) | 1 (33.3) |
| Test-negative case-control | 73 (39) | 27 (51.9) | 7 (63.6) | 2 (66.7) |
| Target trial emulation | 5 (2.7) | 0 (0) | 0 (0) | 0 (0) |
| Quasi-experimental | 2 (1.1) | 1 (1.9) | 0 (0) | 0 (0) |
| Case-control | 11 (5.9) | 3 (5.8) | 2 (18.2) | 0 (0) |
| Ecological study | 1 (0.5) | 0 (0) | 0 (0) | 0 (0) |
| Cross-sectional | 0 (0) | 1 (1.9) | 0 (0) | 0 (0) |
| Study type |  |  |  |  |
| Retrospective | 164 (87.7) | 43 (82.7) | 9 (81.8) | 3 (100) |
| Prospective | 19 (10.2) | 7 (13.5) | 2 (18.2) | 0 (0) |
| Ambispective | 4 (2.1) | 2 (3.8) | 0 (0) | 0 (0) |
| Estimation method* |  |  |  |  |
| Logistic regression | 95 (49.0) | 39 (72.2) | 11 (100.0) | 2 (66.7) |
| Cox proportional hazards | 59 (30.4) | 6 (11.1) | 0 (0) | 0 (0) |
| Poisson regression | 10 (5.2) | 2 (3.7) | 0 (0) | 1 (33.3) |
| Crude statistics | 9 (4.6) | 2 (3.7) | 0 (0) | 0 (0) |
| Kaplan–Meier | 7 (3.6) | 2 (3.7) | 0 (0) | 0 (0) |
| Negative binomial | 7 (3.6) | 0 (0) | 0 (0) | 0 (0) |
| Log-binomial regression | 4 (2.1) | 2 (3.7) | 0 (0) | 0 (0) |
| Regression discontinuity design | 2 (1.0) | 1 (1.9) | 0 (0) | 0 (0) |
| Proportional incidence rate model | 1 (0.5) | 0 (0) | 0 (0) | 0 (0) |
| Comparator group* |  |  |  |  |
| Baseline VE estimates |  |  |  |  |
| Any vaccinated vs unvaccinated | 27 (7.5) | 8 (7.9) | 3 (15.8) | 0 (0) |
| One dose vs unvaccinated | 82 (22.8) | 28 (27.7) | 7 (36.8) | 1 (16.7) |
| Primary vaccination series vs unvaccinated | 118 (32.8) | 36 (35.6) | 9 (47.4) | 2 (33.3) |
| 1st booster dose vs unvaccinated | 63 (17.5) | 16 (15.8) | 0 (0) | 1 (16.7) |
| 2nd booster dose vs unvaccinated | 13 (3.6) | 0 (0) | 0 (0) | 0 (0) |
| 3rd booster dose vs unvaccinated | 1 (0.3) | 0 (0) | 0 (0) | 0 (0) |
| **Dose comparisons** |  |  |  |  |
| First dose vs second dose | 5 (1.4) | 1 (1) | 0 (0) | 1 (16.7) |
| Primary vaccination series vs 1st booster dose | 27 (7.5) | 9 (8.9) | 0 (0) | 1 (16.7) |
| 2nd dose vs 2nd booster dose | 2 (0.6) | 0 (0) | 0 (0) | 0 (0) |
| 1st booster dose vs 2nd booster dose | 8 (2.2) | 0 (0) | 0 (0) | 0 (0) |
| **Booster-specific comparisons** |  |  |  |  |
| Primary vaccination series vs bivalent booster | 1 (0.3) | 0 (0) | 0 (0) | 0 (0) |
| Bivalent booster vs monovalent booster | 2 (0.6) | 0 (0) | 0 (0) | 0 (0) |
| **Vaccine administration comparisons** |  |  |  |  |
| Timing relative to last vaccine dose | 7 (1.9) | 0 (0) | 0 (0) | 0 (0) |
| Different dosages | 1 (0.3) | 1 (1) | 0 (0) | 0 (0) |
| Different vaccine types | 2 (0.6) | 1 (1) | 0 (0) | 0 (0) |
| **Natural immunity comparison** |  |  |  |  |
| Natural infection vs vaccinated | 1 (0.3) | 1 (1) | 0 (0) | 0 (0) |
| **Vaccine type\*** |  |  |  |  |
| All vaccines | 15 (4.6) | 0 (0) | 0 (0) | 0 (0) |
| mRNA vaccines | 28 (8.6) | 1 (0.9) | 0 (0) | 0 (0) |
| Inactivated vaccines | 2 (0.6) | 10 (8.9) | 0 (0) | 0 (0) |
| **Primary series** |  |  |  |  |
| Heterologous schedule | 3 (0.9) | 0 (0) | 0 (0) | 0 (0) |
| AstraZeneca | 41 (12.7) | 18 (16.1) | 7 (28) | 2 (50) |
| BBIBP-CorV (Sinopharm) | 4 (1.2) | 12 (10.7) | 6 (24) | 0 (0) |
| CanSino (Ad5-nCoV) | 0 (0) | 5 (4.5) | 0 (0) | 0 (0) |
| CoronaVac | 11 (3.4) | 27 (24.1) | 1 (4) | 0 (0) |
| Covaxin | 0 (0) | 0 (0) | 3 (12) | 0 (0) |
| CoviVac | 1 (0.3) | 0 (0) | 0 (0) | 0 (0) |
| EpiVacCorona | 1 (0.3) | 0 (0) | 0 (0) | 0 (0) |
| Hayat-Vax | 0 (0) | 1 (0.9) | 0 (0) | 0 (0) |
| Janssen | 15 (4.6) | 4 (3.6) | 1 (4) | 0 (0) |
| Moderna | 65 (20.1) | 3 (2.7) | 3 (12) | 0 (0) |
| Novavax | 1 (0.3) | 0 (0) | 0 (0) | 0 (0) |
| Pfizer-BioNTech | 111 (34.3) | 20 (17.9) | 3 (12) | 2 (50) |
| QazVac | 0 (0) | 1 (0.9) | 0 (0) | 0 (0) |
| Soberana | 0 (0) | 1 (0.9) | 0 (0) | 0 (0) |
| Sputnik V | 4 (1.2) | 4 (3.6) | 1 (4) | 0 (0) |
| WIBP-CorV (Sinopharm) | 0 (0) | 1 (0.9) | 0 (0) | 0 (0) |
| **Booster** |  |  |  |  |
| Monovalent booster (Others, unspecified) | 1 (0.3) | 0 (0) | 0 (0) | 0 (0) |
| Monovalent booster (mRNA, unspecified) | 7 (2.2) | 0 (0) | 0 (0) | 0 (0) |
| Monovalent booster (Pfizer) | 1 (0.3) | 2 (1.8) | 0 (0) | 0 (0) |
| Monovalent booster (Moderna) | 2 (0.6) | 0 (0) | 0 (0) | 0 (0) |
| CoronaVac (booster) | 0 (0) | 2 (1.8) | 0 (0) | 0 (0) |
| Bivalent booster (mRNA) | 7 (2.2) | 0 (0) | 0 (0) | 0 (0) |
| Bivalent booster (Moderna) | 2 (0.6) | 0 (0) | 0 (0) | 0 (0) |
| Bivalent booster (Pfizer) | 2 (0.6) | 0 (0) | 0 (0) | 0 (0) |
| **Matching/Weighting method\*,†** | **83** | **17** | **4** | **3** |
| **No weighting/matching\*\*** | **111 (59.4)** | **35 (67.3)** | **7 (63.6)** | **2 (66.7)** |
| **Matching** |  |  |  |  |
| Exact matching | 55 (66.3) | 13 (76.5) | 3 (75.0) | 1 (100.0) |
| Propensity score matching | 13 (15.7) | 3 (17.5) | 0 (0) | 0 (0) |
| Incidence density sampling | 1 (1.2) | 0 (0) | 1 (25.0) | 0 (0) |
| Random/probabilistic matching | 1 (1.2) | 1 (5.9) | 0 (0) | 0 (0) |
| Rolling entry matching | 1 (1.2) | 0 (0) | 0 (0) | 0 (0) |
| **Weighting** |  |  |  |  |
| IPTW | 7 (8.4) | 0 (0) | 0 (0) | 0 (0) |
| Other weighting methods | 3 (3.6) | 0 (0) | 0 (0) | 0 (0) |
| Overlap weighting | 1 (1.2) | 0 (0) | 0 (0) | 0 (0) |
| sIPT | 1 (1.2) | 0 (0) | 0 (0) | 0 (0) |
| **Covariates matched/weighted/adjusted for\*,†** |  |  |  |  |
| **None described\*\*** | **3 (1.6)** | **2 (3.8)** | **2 (1.8)** | **0 (0)** |
| Demographics | 182 (20.1) | 49 (32.7) | 9 (26.5) | 3 (4.8) |
| Temporal | 133 (14.7) | 29 (19.3) | 5 (14.7) | 3 (4.8) |
| Clinical comorbidities & health status | 125 (13.8) | 37 (24.7) | 3 (8.8) | 3 (4.8) |
| Geography & residence | 117 (12.9) | 18 (12.0) | 1 (2.9) | 3 (4.8) |
| Socioeconomic status | 78 (8.6) | 2 (1.3) | 3 (8.8) | 14 (22.2) |
| Prior SARS-CoV-2 & vaccination history | 66 (7.3) | 0 (0.0) | 1 (2.9) | 13 (20.6) |
| Healthcare utilisation | 48 (5.3) | 6 (4.0) | 3 (8.8) | 1 (1.6) |
| Exposure risks | 42 (4.6) | 2 (1.3) | 1 (2.9) | 13 (20.6) |
| Household & living conditions | 36 (4.0) | 1 (0.7) | 1 (2.9) | 0 (0.0) |
| Testing behaviours and prior infections | 31 (3.4) | 2 (1.3) | 3 (8.8) | 9 (14.3) |
| Physiologic | 29 (3.2) | 1 (0.7) | 1 (2.9) | 1 (1.6) |
| Behavioural & exposure factors | 13 (1.4) | 3 (2.0) | 3 (8.8) | 0 (0.0) |
| Propensity scores | 6 (0.7) | 0 (0.0) | 0 (0.0) | 0 (0.0) |
| **Sensitivity analyses\*,†** |  |  |  |  |
| **None described\*\*** | **65 (34.8)** | **24 (46.2)** | **9 (81.8)** | **0 (0)** |
| Changes to study population inclusion/exclusion criteria | 55 (26.2) | 7 (16.7) | 0 (0.0) | 1 (16.7) |
| Alternate definitions/classifications of exposure | 27 (12.9) | 8 (19.0) | 1 (33.3) | 0 (0.0) |
| Alternate outcome definitions or ascertainment methods | 31 (14.8) | 4 (9.5) | 0 (0.0) | 0 (0.0) |
| Alternate model specifications/statistical approaches | 27 (12.9) | 5 (11.9) | 0 (0.0) | 1 (16.7) |
| Alternate control selection and/or matching methodologies | 17 (8.1) | 7 (16.7) | 1 (33.3) | 0 (0.0) |
| Adjustment for/restriction to specific calendar periods or trends | 16 (7.6) | 2 (4.8) | 0 (0.0) | 2 (33.3) |
| Negative controls | 18 (8.6) | 2 (4.8) | 0 (0.0) | 0 (0.0) |
| Assessment of unmeasured confounding | 9 (4.3) | 3 (7.1) | 0 (0.0) | 0 (0.0) |
| Imputation/assumptions for missing vaccination data | 6 (2.9) | 2 (4.8) | 1 (33.3) | 1 (16.7) |
| Alternate reference/comparator groups | 4 (1.9) | 2 (4.8) | 0 (0.0) | 1 (16.7) |
| **Outcome\*** |  |  |  |  |
| **Infection & reinfection** |  |  |  |  |
| Case status (infection) | 111 (29) | 33 (33.7) | 6 (28.6) | 2 (28.6) |
| Incidence | 1 (0.3) | 0 (0) | 0 (0) | 0 (0) |
| Reinfections | 1 (0.3) | 0 (0) | 0 (0) | 0 (0) |
| **Disease severity & clinical course** |  |  |  |  |
| Mortality | 65 (17) | 16 (16.3) | 4 (19) | 2 (28.6) |
| Case status (severity) | 39 (10.2) | 17 (17.3) | 4 (19) | 1 (14.3) |
| Pneumonia/SARI | 1 (0.3) | 6 (6.1) | 0 (0) | 0 (0) |
| COVID symptoms | 5 (1.3) | 0 (0) | 0 (0) | 0 (0) |
| Disease duration | 2 (0.5) | 0 (0) | 0 (0) | 0 (0) |
| COVID-19 sick leave | 1 (0.3) | 0 (0) | 0 (0) | 0 (0) |
| Post-acute sequelae | 1 (0.3) | 0 (0) | 0 (0) | 0 (0) |
| **Healthcare utilisation** |  |  |  |  |
| Hospitalisation | 102 (26.6) | 16 (16.3) | 5 (23.8) | 2 (28.6) |
| ICU admission | 22 (5.7) | 2 (2) | 2 (9.5) | 0 (0) |
| ED attendance | 15 (3.9) | 0 (0) | 0 (0) | 0 (0) |
| Receipt of life-support interventions | 13 (3.4) | 2 (2) | 0 (0) | 0 (0) |
| **Virological & immunological markers** |  |  |  |  |
| Neutralising antibody levels | 0 (0) | 2 (2) | 0 (0) | 0 (0) |
| Viral shedding duration | 0 (0) | 1 (1) | 0 (0) | 0 (0) |
| **Transmission dynamics** |  |  |  |  |
| Transmission (including secondary attacks) | 1 (0.3) | 3 (3) | 0 (0) | 0 (0) |

On study designs, LMIC studies used test-negative case-control designs most often (7/11, 63.6%) and cohort designs less frequently (2/11, 18.2%); HIC balanced both (cohort 95/187, 50.8%; test-negative 73/187, 39.0%), and UMIC leaned toward test-negative designs (27/52, 51.9%). Estimator use was overwhelmingly logistic regression in LMIC (11/11, 100.0%), common in UMIC (39/54, 72.2%), and less dominant in HIC (95/194, 49.0%), while Cox regression models were concentrated in HIC studies (59/194, 30.4%) and rare elsewhere (UMIC 6/54, 11.1%; LMIC 0/11). Within the vaccine-product domain, HIC studies most often assessed Pfizer-BioNTech (111, 34.3%) and Moderna (65, 20.1%), whereas UMIC emphasised CoronaVac (27, 24.1%) and Pfizer-BioNTech (20, 17.9%); LMIC studies frequently assessed AstraZeneca (7, 28.0%) and BBIBP-CorV (6, 24.0%). Booster and bivalent comparisons were largely confined to HIC studies (e.g., first-booster vs unvaccinated: 63, 17.5%; bivalent vs monovalent: 2, 0.6%), with no booster-vs-unvaccinated contrasts in LMIC.

Most studies reported no matching/weighting (HIC 111/187, 59.4%; UMIC 35/52, 67.3%; LMIC 7/11, 63.6%; multinational 2/3, 66.7%). Among analyses using these tools (domain denominators: HIC n=83, UMIC n=17, LMIC n=4, multinational n=1), exact matching (including coarsened exact matching) predominated (HIC 66.3%; UMIC 76.5%; LMIC 75.0%; multinational 100%), with IPTW was used only in HIC (8.4%). Very few studies omitted covariate adjustment entirely (HIC 3/187, 1.6%; UMIC 2/52, 3.8%; LMIC 2/11, 18.2%; multinational 0/3, 0). Sensitivity analyses were least common in LMIC studies (none described in 9/11, 81.8%), compared with HIC (65/187, 34.8%) and UMIC (24/52, 46.2%). Outcome emphasis differed modestly: infection was most frequently analysed across strata (HIC 111, 29.0%; UMIC 33, 33.7%; LMIC 6, 28.6%), with hospitalisation (HIC 102, 26.6%; UMIC 16, 16.3%; LMIC 5, 23.8%) and mortality (HIC 65, 17.0%; UMIC 16, 16.3%; LMIC 4, 19.0%) next most common.

Stratified by year (**Table 4**), the geographic mix shifted from HIC-dominant (28/33, 84.8% in 2021) toward greater UMIC representation (11/35, 31.4% in 2024). Over the same period, publication lag lengthened sharply—from 141 days (IQR 107–178) in 2021 to 673 (IQR 548–863) in 2024—while median individual-level cohort size fell from 156 930 (IQR 15 101–982 005) to 19 314 (IQR 6 864–769 448). Comparator mixes moved away from primary-series vs unvaccinated (38.7% to 33.8%) and one-dose vs unvaccinated (41.9% to 20.0%), toward first-booster vs unvaccinated (0% to 16.9) and primary-series vs first-booster (1.6% to 10.8%). Vaccine portfolios in 2021–2022 were led by Pfizer-BioNTech and Moderna, broadening to include bivalents only in 2023–2024. The share reporting no sensitivity analyses was 12/33 (36.4%) in 2021 and 16/35 (45.7%) in 2024. Across years, infection, hospitalisation, and mortality remained the leading endpoints (infection: 28.2–31.9%; hospitalisation: 21.5–30.4%; mortality: 13.8–23.2%), with transmission and immunologic markers consistently rare. Target-trial emulation as a study design first appeared in 2022 (1/113, 0.9%) and rose to 2/35 (5.7%) in 2024. Within the estimation-method domain, logistic regression remained predominant (55.9%, 56.4%, 57.3%, 52.8% in 2021– 2024), while the shares using Cox and Poisson increased (Cox 17.6% to 33.3%; Poisson 2.9% to 8.3%) and Kaplan–Meier and negative binomial fell to 0% by 2024. Studies with no matching/weighting comprised the majority each year, with 18/33 (54.5%), 74/113 (65.5%), 43/72 (59.7%), and 20/35 (57.1%) in 2021, 2022, 2023, and 2024, respectively. Among analyses that did match/weight, exact matching remained the most common, and usage of propensity-score matching increased from 12.5% (2021) to 31.2% (2024). Covariate adjustment tightened over time: studies with no adjustment fell from 3/33 (9.1%) in 2021 to 0/35 (0%) in 2024, with demographics (∼20% each year) and calendar time (∼14–15%) being the dominant factors adjusted for. Additionally, clinical comorbidity adjustment rose to 17.1% by 2024. Accounting for prior immunity/vaccination increased after 2021 (3.4% to 7.3%), but adjustment for testing behaviour, socioeconomic, and behavioural factors is sparse from 2021–2024 (≈1–9%).

**Table 4.** Characteristics of included studies by publication year. Values are n (%) within each publication-year stratum unless stated; medians are reported with interquartile ranges (IQR). Column denominators (studies per year): 2021, N=33; 2022, N=113; 2023, N=72; 2024, N=35. Definitions for study-design categories, estimation methods, comparator mappings, vaccine product labels, and outcome groupings follow Table 2. Abbreviations: VE, vaccine effectiveness; IQR, interquartile range; ED, emergency department; ICU, intensive care unit; SARI, severe acute respiratory infection; IPTW, inverse probability of treatment weighting; sIPT, stabilised inverse probability of treatment weights. * denotes multi-select domains. These use the domain-specific observation total within each year stratum as the denominator; studies may contribute >1 observation; minor deviations reflect rounding. ^†^ Rows “No weighting/matching,” “None” (covariates), and “None described” (sensitivity analyses) use per-year study counts: 2021 N=33; 2022 N=113; 2023 N=72; 2024 N=35. All other rows use within-section totals excluding the “None/No …” row: **Matching/Weighting** n=16, 40, 33, 16 (2021–2024); **Covariates** n=145, 499, 341, 163; **Sensitivity analyses** n=37, 116, 76, 32. (N = studies; n = within-section total.)

| Category | Publication year |  |  |  |
| --- | --- | --- | --- | --- |
|  | 2021<br>(N = 33) | 2022<br>(N = 113) | 2023<br>(N = 72) | 2024<br>(N = 35) |
| Publication year |  |  |  |  |
| High income | 28 (84.8) | 84 (74.3) | 52 (72.2) | 23 (65.7) |
| Upper middle income | 3 (9.1) | 19 (16.8) | 19 (26.4) | 11 (31.4) |
| Lower middle income | 2 (6.1) | 8 (7.1) | 0 (0) | 1 (2.9) |
| Multinational | 0 (0) | 2 (1.8) | 1 (1.4) | 0 (0) |
| Median time to publication |  |  |  |  |
|  | 141 (107–178) | 193 (116–305) | 373 (257–544) | 673 (548–863) |
| Cohort size (median [IQR]) |  |  |  |  |
| Individuals | 156930 (15101–982005) | 195276 (10885–1527603) | 122432 (12189–517335) | 19314 (6864–769448) |
| Households | 173569 (173569–173569) | N/A | 20651 (20651–20651) | N/A |
| Tests | 75546 (75546–75546) | 180016 (31160–478455) | 1718284 (946552–2490017) | 34412 (27205–41619) |
| Study type |  |  |  |  |
| Cohort | 16 (48.5) | 49 (43.4) | 35 (48.6) | 18 (51.4) |
| Test-negative case-control | 15 (45.5) | 54 (47.8) | 27 (37.5) | 13 (37.1) |
| Target trial emulation | 0 (0) | 1 (0.9) | 2 (2.8) | 2 (5.7) |
| Quasi-experimental | 1 (3.0) | 1 (0.9) | 0 (0) | 1 (2.9) |
| Case-control | 1 (3.0) | 7 (6.2) | 7 (9.7) | 1 (2.9) |
| Ecological study | 0 (0) | 0 (0) | 1 (1.4) | 0 (0) |
| Cross-sectional | 0 (0) | 1 (0.9) | 0 (0) | 0 (0) |
| Study type |  |  |  |  |
| Retrospective | 29 (87.9) | 97 (85.8) | 64 (88.9) | 29 (82.9) |
| Prospective | 4 (12.1) | 13 (11.5) | 7 (9.7) | 4 (11.4) |
| Ambispective | 0 (0) | 3 (2.7) | 1 (1.4) | 2 (5.7) |
| Estimation method* |  |  |  |  |
| Logistic regression | 19 (55.9) | 66 (56.4) | 43 (57.3) | 19 (52.8) |
| Cox proportional hazards | 6 (17.6) | 29 (24.8) | 18 (24.0) | 12 (33.3) |
| Poisson regression | 1 (2.9) | 5 (4.3) | 4 (5.3) | 3 (8.3) |
| Crude statistics | 2 (5.9) | 6 (5.1) | 2 (2.7) | 1 (2.8) |
| Kaplan–Meier | 1 (2.9) | 5 (4.3) | 3 (4.0) | 0 (0) |
| Negative binomial | 3 (8.8) | 3 (2.6) | 1 (1.3) | 0 (0) |
| Log-binomial regression | 1 (2.9) | 2 (1.7) | 3 (4.0) | 0 (0) |
| Regression discontinuity design | 1 (2.9) | 1 (0.9) | 0 (0) | 1 (2.8) |
| Proportional incidence rate model | 0 (0) | 0 (0) | 1 (1.3) | 0 (0) |
| Comparator group* |  |  |  |  |
| Baseline VE estimates |  |  |  |  |
| Any vaccinated vs unvaccinated | 7 (11.3) | 21 (10.1) | 7 (4.6) | 3 (4.6) |
| One dose vs unvaccinated | 26 (41.9) | 55 (26.4) | 24 (15.9) | 13 (20) |
| Primary vaccination series vs unvaccinated | 24 (38.7) | 76 (36.5) | 43 (28.5) | 22 (33.8) |
| 1st booster dose vs unvaccinated | 0 (0) | 33 (15.9) | 36 (23.8) | 11 (16.9) |
| 2nd booster dose vs unvaccinated | 0 (0) | 3 (1.4) | 7 (4.6) | 3 (4.6) |
| 3rd booster dose vs unvaccinated | 0 (0) | 0 (0) | 0 (0) | 1 (1.5) |
| **Dose comparisons** |  |  |  |  |
| First dose vs second dose | 3 (4.8) | 3 (1.4) | 1 (0.7) | 0 (0) |
| Primary vaccination series vs 1st booster | 1 (1.6) | 11 (5.3) | 18 (11.9) | 7 (10.8) |
| 2nd dose vs 2nd booster dose | 0 (0) | 0 (0) | 2 (1.3) | 0 (0) |
| 1st booster dose vs 2nd booster dose | 0 (0) | 1 (0.5) | 7 (4.6) | 0 (0) |
| **Booster-specific comparisons** |  |  |  |  |
| Primary series vs bivalent booster | 0 (0) | 0 (0) | 1 (0.7) | 0 (0) |
| Bivalent booster vs monovalent booster | 0 (0) | 0 (0) | 1 (0.7) | 1 (1.5) |
| **Vaccine administration comparisons** |  |  |  |  |
| Timing relative to last vaccine dose | 1 (1.6) | 2 (1.0) | 2 (1.3) | 2 (3.1) |
| Different dosages | 0 (0) | 0 (0) | 0 (0) | 2 (3.1) |
| Different vaccine types | 0 (0) | 1 (0.5) | 2 (1.3) | 0 (0) |
| **Natural immunity comparison** |  |  |  |  |
| Natural infection vs vaccinated | 0 (0) | 2 (1.0) | 0 (0) | 0 (0) |
| **Vaccine type\*** |  |  |  |  |
| All vaccines | 2 (3.7) | 4 (1.8) | 4 (2.9) | 5 (8.9) |
| mRNA vaccines | 3 (5.6) | 11 (5.0) | 11 (8.1) | 4 (7.1) |
| Inactivated vaccines | 1 (1.9) | 3 (1.4) | 5 (3.7) | 3 (5.4) |
| **Primary series** |  |  |  |  |
| Heterologous schedule | 0 (0) | 2 (0.9) | 0 (0) | 1 (1.8) |
| AstraZeneca | 10 (18.5) | 32 (14.6) | 18 (13.2) | 8 (14.3) |
| BBIBP-CorV (Sinopharm) | 1 (1.9) | 13 (5.9) | 4 (2.9) | 4 (7.1) |
| CanSino (Ad5-nCoV) | 0 (0) | 2 (0.9) | 3 (2.2) | 0 (0) |
| CoronaVac | 1 (1.9) | 15 (6.8) | 17 (12.5) | 6 (10.7) |
| Covaxin | 1 (1.9) | 2 (0.9) | 0 (0) | 0 (0) |
| CoviVac | 0 (0) | 1 (0.5) | 0 (0) | 0 (0) |
| EpiVacCorona | 0 (0) | 1 (0.5) | 0 (0) | 0 (0) |
| Hayat-Vax | 0 (0) | 0 (0) | 1 (0.7) | 0 (0) |
| Janssen | 2 (3.7) | 13 (5.9) | 4 (2.9) | 1 (1.8) |
| Moderna | 10 (18.5) | 40 (18.3) | 16 (11.8) | 5 (8.9) |
| Novavax | 0 (0) | 0 (0) | 1 (0.7) | 0 (0) |
| Pfizer-BioNTech | 21 (38.9) | 70 (32.0) | 32 (23.5) | 13 (23.2) |
| QazVac | 0 (0) | 0 (0) | 1 (0.7) | 0 (0) |
| Soberana | 0 (0) | 0 (0) | 0 (0) | 1 (1.8) |
| Sputnik V | 2 (3.7) | 3 (1.4) | 2 (1.5) | 2 (3.6) |
| WIBP-CorV (Sinopharm) | 0 (0) | 0 (0) | 1 (0.7) | 0 (0) |
| **Booster** |  |  |  |  |
| Monovalent booster (Others, unspecified) | 0 (0) | 0 (0) | 1 (0.7) | 0 (0) |
| Monovalent booster (mRNA, unspecified) | 0 (0) | 0 (0) | 5 (3.7) | 2 (3.6) |
| Monovalent booster (Pfizer) | 0 (0) | 3 (1.4) | 0 (0) | 0 (0) |
| Monovalent booster (Moderna) | 0 (0) | 2 (0.9) | 0 (0) | 0 (0) |
| CoronaVac (booster) | 0 (0) | 2 (0.9) | 0 (0) | 0 (0) |
| Bivalent booster (mRNA) | 0 (0) | 0 (0) | 6 (4.4) | 1 (1.8) |
| Bivalent booster (Moderna) | 0 (0) | 0 (0) | 2 (1.5) | 0 (0) |
| Bivalent booster (Pfizer) | 0 (0) | 0 (0) | 2 (1.5) | 0 (0) |
| **Matching/Weighting method\*<sup>†</sup>** |  |  |  |  |
| **No weighting/matching** | 18 (54.5) | 74 (65.5) | 43 (59.7) | 20 (57.1) |
| **Matching** |  |  |  |  |
| Exact matching | 12 (75.0) | 31 (77.5) | 21 (63.6) | 8 (50.0) |
| Propensity score matching | 2 (12.5) | 3 (7.5) | 6 (18.2) | 5 (31.2) |
| Incidence density sampling | 0 (0.0) | 2 (5.0) | 0 (0.0) | 0 (0.0) |
| Random/probabilistic matching | 0 (0.0) | 0 (0.0) | 1 (3.0) | 1 (6.2) |
| Rolling entry matching | 0 (0.0) | 1 (2.5) | 0 (0.0) | 0 (0.0) |
| **Weighting** |  |  |  |  |
| IPTW | 1 (6.2) | 2 (5.0) | 4 (12.1) | 0 (0.0) |
| Other weighting methods | 1 (6.2) | 1 (2.5) | 0 (0.0) | 1 (6.2) |
| Overlap weighting | 0 (0.0) | 0 (0.0) | 1 (3.0) | 0 (0.0) |
| sIPT | 0 (0.0) | 0 (0.0) | 0 (0.0) | 1 (6.2) |
| **Covariates matched/weighted/adjusted for\*<sup>†</sup>** |  |  |  |  |
| **None described** | 3 (9.1) | 2 (1.8) | 2 (2.8) | 0 (0.0) |
| Demographics | 32 (21.9) | 108 (21.4) | 68 (19.9) | 34 (20.7) |
| Temporal | 20 (13.7) | 76 (15.1) | 50 (14.6) | 23 (14.0) |
| Clinical comorbidities & health status | 18 (12.3) | 70 (13.9) | 51 (14.9) | 28 (17.1) |
| Geography & residence | 15 (10.3) | 71 (14.1) | 36 (10.5) | 17 (10.4) |
| Socioeconomic status | 15 (10.3) | 42 (8.3) | 27 (7.9) | 13 (7.9) |
| Prior SARS-CoV-2 & vaccination history | 5 (3.4) | 31 (6.2) | 31 (9.1) | 12 (7.3) |
| Healthcare utilisation | 10 (6.8) | 23 (4.6) | 15 (4.4) | 10 (6.1) |
| Exposure risks | 6 (4.1) | 19 (3.8) | 24 (7.0) | 8 (4.9) |
| Household & living conditions | 7 (4.8) | 15 (3.0) | 14 (4.1) | 2 (1.2) |
| Testing behaviours and prior infections | 9 (6.2) | 24 (4.8) | 8 (2.3) | 4 (2.4) |
| Physiologic | 4 (2.7) | 13 (2.6) | 7 (2.0) | 8 (4.9) |
| Behavioural & exposure factors | 3 (2.1) | 6 (1.2) | 7 (2.0) | 3 (1.8) |
| Propensity scores (as covariate) | 1 (0.7) | 1 (0.2) | 3 (0.9) | 1 (0.6) |
| **Sensitivity analyses\*<sup>†</sup>** |  |  |  |  |
| None described | 12 (36.4) | 43 (38.1) | 27 (37.5) | 16 (45.7) |
| Changes to study population inclusion/exclusion criteria | 7 (18.9) | 26 (22.4) | 22 (28.9) | 8 (25.0) |
| Alternate definitions/classifications of exposure | 4 (10.8) | 18 (15.5) | 8 (10.5) | 6 (18.8) |
| Alternate outcome definitions or ascertainment methods | 5 (13.5) | 14 (12.1) | 13 (17.1) | 3 (9.4) |
| Alternate model specifications/statistical approaches | 7 (18.9) | 12 (10.3) | 11 (14.5) | 3 (9.4) |
| Alternate control selection and/or matching methodologies | 4 (10.8) | 13 (11.2) | 6 (7.9) | 2 (6.2) |
| Adjustment for or restriction to specific calendar periods or temporal trends | 5 (13.5) | 6 (5.2) | 5 (6.6) | 4 (12.5) |
| Negative controls | 2 (5.4) | 9 (7.8) | 4 (5.3) | 5 (15.6) |
| Assessment of unmeasured confounding | 2 (5.4) | 6 (5.2) | 4 (5.3) | 0 (0.0) |
| Imputation or different assumptions for handling missing vaccination data | 0 (0.0) | 7 (6.0) | 2 (2.6) | 1 (3.1) |
| Alternate reference/comparator groups | 1 (2.7) | 5 (4.3) | 1 (1.3) | 0 (0.0) |
| **Outcome\*** |  |  |  |  |
| **Infection & reinfection** |  |  |  |  |
| Case status (infection) | 22 (31.9) | 73 (29.9) | 37 (28.2) | 20 (30.8) |
| Incidence | 0 (0) | 0 (0) | 0 (0) | 1 (1.5) |
| Reinfections | 0 (0) | 0 (0) | 0 (0) | 1 (1.5) |
| **Disease severity & clinical course** |  |  |  |  |
| Mortality | 16 (23.2) | 41 (16.8) | 21 (16.0) | 9 (13.8) |
| Case status (severity) | 4 (5.8) | 28 (11.5) | 19 (14.5) | 10 (15.4) |
| Pneumonia/SARI | 0 (0) | 3 (1.2) | 3 (2.3) | 1 (1.5) |
| COVID symptoms | 0 (0) | 3 (1.2) | 1 (0.8) | 1 (1.5) |
| Disease duration (symptoms/isolation) | 0 (0) | 0 (0) | 2 (1.5) | 0 (0) |
| COVID-19 sick leave | 0 (0) | 0 (0) | 0 (0) | 1 (1.5) |
| Post-acute sequelae | 0 (0) | 0 (0) | 1 (0.8) | 0 (0) |
| **Healthcare utilisation** |  |  |  |  |
| Hospitalisation | 21 (30.4) | 57 (23.4) | 33 (25.2) | 14 (21.5) |
| ICU admission | 4 (5.8) | 15 (6.1) | 5 (3.8) | 2 (3.1) |
| ED attendance | 2 (2.9) | 7 (2.9) | 3 (2.3) | 3 (4.6) |
| Receipt of life-support interventions | 0 (0) | 11 (4.5) | 3 (2.3) | 1 (1.5) |
| **Virological & immunological markers** |  |  |  |  |
| Neutralising antibody levels | 0 (0) | 2 (0.8) | 0 (0) | 0 (0) |
| Viral shedding duration | 0 (0) | 1 (0.4) | 0 (0) | 0 (0) |
| **Transmission dynamics** |  |  |  |  |
| Transmission (including secondary attacks) | 0 (0) | 0 (0) | 3 (2.3) | 1 (1.5) |

There was a clustering of analytic approaches around a few familiar methods to obtain VE estimates (**Table 5**). Almost every test-negative study used logistic regression (106/109, 97.2%), whereas cohort studies split across time-to-event modelling [Cox regression: 60/118, 48.0%; Kaplan–Meier (7/118, 5.6%)], count models (Poisson + negative binomial regression: 18/118, 15.3%), and logistic models (26/118, 20.8%); log-binomial regression (6/118, 4.8%), and crude estimates (8/118, 6.4%) were less common. Articles employing a case-control design leaned heavily on logistic regression (15/16, 83.3%). Quasi-experimental analyses all employed a regression discontinuity design (3/3, 100%), while the single cross-sectional study reported crude statistics and an ecological study used a proportional incidence rate model.

**Table 5.** Analytic methods, sensitivity analyses, and covariate adjustment by study design. Values are n (%) within each study-design stratum unless stated. Column denominators (number of studies per stratum): Cohort n = 118; Test-negative case-control n = 109; Case-control n = 16; Target-trial emulation n = 5; Quasi-experimental n = 3; Cross-sectional n = 1; Ecological n = 1. Percentages in strata with very small n (e.g., target-trial emulation, quasi-experimental, cross-sectional, ecological) should be interpreted cautiously. * denotes multi-select domains. These use the domain-specific observation total within each study-design stratum as the denominator; studies may contribute >1 observation; minor deviations reflect rounding. ^†^ Rows “None described” (sensitivity analyses/validation) and “None” (covariates) use the study counts in each design stratum: Cohort N=118; Test-negative case-control N=109; Case-control N=16; Target-trial emulation N=5; Quasi-experimental N=3; Cross-sectional N=1; Ecological study N=1. All other rows in multi-contribution domains use the design-specific observation totals (a study may contribute >1 observation within a domain). **Sensitivity analyses (excluding “Not described**”): Cohort n=101, Test-negative n=130, Case-control n=10, Target-trial emulation n=10, Quasi-experimental n=7, Cross-sectional n=1, Ecological n=2. **Covariates weighted/matched/adjusted for (excluding “None”)**: Cohort n=526, Test-negative n=520, Case-control n=72, Target-trial emulation n=29, Quasi-experimental n=5, Cross-sectional n=1, Ecological n=

|  | Cohort<br>(n = 118) | Test-negative case-control<br>(n = 109) | Case-control<br>(n = 16) | Study design<br>Target trial<br>emulation<br>(n = 5) | Quasi-experimental<br>(n = 3) | Cross-sectional<br>(n = 1) | Ecological<br>study<br>(n = 1) |
| --- | --- | --- | --- | --- | --- | --- | --- |
| Estimation method |  |  |  |  |  |  |  |
| Cox proportional hazards | 60 (48) | 0 (0) | 2 (11.1) | 3 (60) | 0 (0) | 0 (0) | 0 (0) |
| Kaplan-Meier | 7 (5.6) | 0 (0) | 1 (5.6) | 1 (20) | 0 (0) | 0 (0) | 0 (0) |
| Logistic regression | 26 (20.8) | 106 (97.2) | 15 (83.3) | 0 (0) | 0 (0) | 0 (0) | 0 (0) |
| Log-binomial regression | 6 (4.8) | 0 (0) | 0 (0) | 0 (0) | 0 (0) | 0 (0) | 0 (0) |
| Negative binomial | 7 (5.6) | 0 (0) | 0 (0) | 0 (0) | 0 (0) | 0 (0) | 0 (0) |
| Poisson regression | 11 (8.8) | 1 (0.9) | 0 (0) | 1 (20) | 0 (0) | 0 (0) | 0 (0) |
| Crude statistics | 8 (6.4) | 2 (1.8) | 0 (0) | 0 (0) | 0 (0) | 1 (100) | 0 (0) |
| Proportional incidence rate model | 0 (0) | 0 (0) | 0 (0) | 0 (0) | 0 (0) | 0 (0) | 1 (100) |
| Regression discontinuity design | 0 (0) | 0 (0) | 0 (0) | 0 (0) | 3 (100) | 0 (0) | 0 (0) |
| Sensitivity analyses*.<sup>†</sup> |  |  |  |  |  |  |  |
| Not described | 60 (50.8) | 29 (26.6) | 8 (50) | 0 (0) | 1 (33.3) | 0 (0) | 0 (0) |
| Alternate control selection and/or matching methodologies | 6 (5.9) | 17 (13.1) | 2 (20) | 0 (0) | 0 (0) | 0 (0) | 0 (0) |
| Alternate definitions/classifications of exposure | 20 (19.8) | 13 (10.0) | 1 (10) | 0 (0) | 2 (28.6) | 0 (0) | 0 (0) |
| Alternate model specifications/statistical approaches | 19 (18.8) | 10 (7.7) | 2 (20) | 0 (0) | 1 (14.2) | 0 (0) | 1 (50) |
| Alternate outcome definitions or ascertainment methods | 12 (11.9) | 19 (14.6) | 3 (30) | 1 (10) | 0 (0) | 0 (0) | 0 (0) |
| Changes to study population inclusion/exclusion criteria | 18 (17.8) | 41 (31.5) | 2 (20) | 2 (20) | 0 (0) | 0 (0) | 0 (0) |
| Adjustment for or restriction to specific calendar periods or temporal trends | 8 (7.9) | 8 (6.2) | 0 (0) | 2 (20) | 2 (28.6) | 0 (0) | 0 (0) |
| Alternate reference/comparator groups | 3 (3.0) | 4 (3.1) | 0 (0) | 0 (0) | 0 (0) | 0 (0) | 0 (0) |
| Assessment of unmeasured confounding | 7 (6.9) | 2 (1.5) | 0 (0) | 1 (10) | 0 (0) | 1 (100) | 1 (50) |
| Imputation and different assumptions for handling missing vaccination data | 3 (3.0) | 7 (5.4) | 0 (0) | 0 (0) | 0 (0) | 0 (0) | 0 (0) |
| Negative controls | 5 (5.0) | 9 (6.9) | 0 (0) | 4 (40) | 2 (28.6) | 0 (0) | 0 (0) |
| Covariates weighted/matched/adjusted for*.<sup>†</sup> |  |  |  |  |  |  |  |
| None** | 4 (3.4) | 2 (1.8) | 1 (6.3) | 0 (0.0) | 1 (33.3) | 0 (0.0) | 0 (0.0) |
| Behavioural & exposure factors | 3 (0.6) | 13 (2.5) | 3 (4.2) | 0 (0.0) | 0 (0.0) | 0 (0.0) | 0 (0.0) |
| Clinical comorbidities & health status | 80 (15.2) | 66 (12.7) | 14 (19.4) | 5 (17.2) | 1 (25) | 0 (0.0) | 1 (33.3) |
| Demographic | 113 (21.5) | 106 (20.4) | 15 (20.8) | 5 (17.2) | 2 (50) | 1 (100.0) | 0 (0.0) |
| Exposure risks | 27 (5.1) | 25 (4.8) | 5 (6.9) | 0 (0.0) | 0 (0.0) | 0 (0.0) | 0 (0.0) |
| Geography & residence | 61 (11.6) | 70 (13.5) | 4 (5.6) | 4 (13.8) | 0 (0.0) | 0 (0.0) | 0 (0.0) |
| Healthcare utilisation | 25 (4.8) | 27 (5.2) | 5 (6.9) | 1 (3.4) | 0 (0.0) | 0 (0.0) | 0 (0.0) |
| Household & living conditions | 19 (3.6) | 18 (3.5) | 1 (1.4) | 0 (0.0) | 0 (0.0) | 0 (0.0) | 0 (0.0) |
| Physiologic | 15 (2.9) | 14 (2.7) | 1 (1.4) | 2 (6.9) | 0 (0.0) | 0 (0.0) | 0 (0.0) |
| Testing behaviours and prior infections | 13 (2.5) | 31 (6.0) | 1 (1.4) | 0 (0.0) | 0 (0.0) | 0 (0.0) | 0 (0.0) |
| Socioeconomic status | 49 (9.3) | 38 (7.3) | 7 (9.7) | 2 (6.9) | 1 (25) | 0 (0.0) | 0 (0.0) |
| Temporal | 68 (12.9) | 83 (16.0) | 12 (16.7) | 5 (17.2) | 0 (0.0) | 0 (0.0) | 1 (33.3) |
| Prior SARS-CoV-2 & vaccination history | 48 (9.1) | 23 (4.4) | 3 (4.2) | 4 (13.8) | 0 (0.0) | 0 (0.0) | 1 (33.3) |
| Propensity scores | 1 (0.2) | 4 (0.8) | 0 (0.0) | 1 (3.4) | 0 (0.0) | 0 (0.0) | 0 (0.0) |

A moderate number of studies probed robustness: about half of cohort reports described no sensitivity analysis (60/118, 50.8%). Negative-control checks and assessments of unmeasured confounding were rare across designs; when present, they were proportionally more common in target-trial emulations (negative controls 4/10, 40.0%; unmeasured confounding 1/10, 10.0%) than in cohorts (5/101, 5.0%; 7/101, 6.9%) or test-negative studies (9/130, 6.9%; 2/130, 1.5%), though absolute counts were small overall. Across designs, adjustment clustered in demographics and clinical comorbidity domains—cohorts: 113 (21.5%) and 80 (15.2%); test-negative: 106 (20.4%) and 66 (12.7%); case-control: 15 (20.8%) and 14 (19.4%). Calendar time was routinely controlled (test-negative 83 [16.0%]; cohorts 68 [12.9%]), and geography/residence was also common (test-negative 70 [13.5%]; cohorts 61 [11.6%]). Covariates closest to infection risk or selection into testing (behaviour/exposure, testing or prior infection, household context, healthcare utilisation) were used less often with generally ≤7% within a design though test-negative studies more often included testing behaviour/prior infection (31 [6.0%]). Use of propensity scores as an adjustment covariate was rare (cohorts 1 [0.2%]; test-negative 4 [0.8%]). Purely unadjusted models were uncommon (e.g., cohorts 4/118, 3.4%; test-negative 2/109, 1.8%).

## Discussion

The present review extends early appraisals of COVID-19 vaccine effectiveness (VE) by tracing how observational methods evolved across 253 studies from 61 countries (2021–2024), highlighting how methodological choices were shaped by available infrastructures, the shifting epidemiological context, and the urgency of decision-making. Before COVID-19, observational VE work, especially for influenza,relied primarily on the test-negative design (TND) which can be easily implemented when data on routine laboratory testing for medically attended acute respiratory illness is available. The TND’s efficiency and its partial control for health-care–seeking bias are well described, as are its identification conditions and pitfalls (calendar-time control, constancy of non-target pathogens across vaccination strata, care-seeking patterns).(21,283,284)

COVID-19 marked a decisive shift in settings with robust registries: individual-level vaccination and testing records made large-scale cohorts feasible, yielding near parity between cohorts assembled from person-level registries (118/253, 46.6%) and TND studies based on routine laboratory databases (109/253, 43.1%). Where linkage remained incomplete—more common in upper-and lower-middle-income settings—TND persisted as the default. Quasi-experimental strategies were rare (3/253, 1.2%), with all employing regression discontinuity design.(6,192,244) This is likely as rollout programmes seldom created sharp, stable discontinuities as eligibility thresholds were introduced and relaxed quickly, rollouts were staggered, and multiple eligibility routes coexisted (e.g., age bands alongside occupational or clinical indications).(17,285,286) Given limited data around cut-off thresholds and frequent cross-overs, there is often insufficient exogenous variation to apply regression-discontinuity designs for estimating VE. Additionally, contrasting with influenza VE literature where instrumental variables (IV) have been used to address unmeasured confounding,(287,288) IV analyses were absent in the COVID-19 VE literature, likely as overlapping, rapidly shifting rollouts made valid instruments difficult to defend.(289)

Analytical choices tracked data infrastructure and surveillance capacity. High-income countries contributed 187/253 (73.9%) of studies, and outcomes that require record linkage—documented infection (152 occurrences across studies), hospitalisation (125 occurrences), and mortality (87 occurrences)—predominated. Publication speed and scale varied by income group: median time to publication was shorter in high-income settings (245 days) than in LMICs (338) and UMICs (391), and median cohort size fell from 178 153 (HIC) to 16 201 (UMIC) and 1 731 (LMIC). Timeliness deteriorated overall, more than quadrupling from a median of 141 days in 2021 to 673 days in 2024, while median individual-level cohort size fell from 156 930 (15 101–982 005) to 19 314 (6 864–769 448). These patterns are consistent with the transition from emergency pipelines to routine reporting, the expiration of temporary data-sharing arrangements, and reductions in PCR testing and genomic sequencing that complicated lineage linkage and variant-specific VE. In the United States, surveillance cadence shifted after the public-health emergency ended;(20) WHO guidance likewise moved toward genomic integration within sentinel systems.(290) Concurrently, sequencing volumes declined sharply in 2023-24, further limiting variant attribution.(291) Nonetheless, this publication lag implies that variant-resolved VE for updated formulations is increasingly generated with substantial delay, risking lagged policy decisions precisely when timely evidence against more immune-evasive lineages is most needed.

This concentration of COVID-19 VE studies in HICs with unequal participation in such studies from LMICs illustrates major disparities in both vaccine access and research capacity across countries. While efforts have been made to strengthen infrastructure in LMICs to support large scale phase-3 clinical trials of SARS-CoV-2 vaccines,(292) existing surveillance and research capacity needs to be continuously maintained and upgraded. For instance, automated surveillance systems for acute-respiratory-infection, enabled through access to electronic-health-records (EHRs), can be pivoted towards real-world vaccine-effectiveness studies during a pandemic scenario.(293) While the COVID-19 pandemic underscored the pivotal role of EHRs in enabling real-world vaccine-effectiveness studies, progress on implementing and using EHRs remains sluggish globally; a 2021 OECD survey found that only 15 countries had a nationally unified EHR.(294) Limitations of electronic data capture for population-based data collection in LMICs include language barriers, unstable internet connection, and overly complicated data collection templates.(295) Additionally, access to diagnostic testing is pivotal in identifying cases of vaccine-breakthrough infection, yet substantial disparities exist in testing capacity, with stark heterogeneities associated with socioeconomic and gender disparities.(296) Improved geographical accessibility to vaccination,(297) and research capacity enhancement pertaining to real-world evaluation of VE is essential to both pandemic preparedness and global health equity.

Estimator choices reflected data availability and pre-pandemic practice. Logistic regression (148/253, 56.1%) and Cox regression (65/253, 24.8%) accounted for ∼81% of estimators with the choice of estimation methods reflecting the availability of data. Early studies with access to person-level data employed person-day Poisson regressions to account for rapid changes in the force of infection;(38,97) with later work employing Cox models with calendar-date time scales to study time-since-vaccination.(42,48,49,56,71,73,75,78,85,92,96,120,121,128–130,132,134,136,138,139,141,144,146,149–152,156,158,160,161,172,190,193,202,215,224,228,231–233,238,241,246,253,258,266,272,275,276,278,298–304) Studies which only had access to hospitalisation or medical attendance data large relied on TND methods. Nearly all TND studies used logistic regression (106/109, 97.2%), while cohorts split across Cox regression (60/118, 48.0%), logistic regression (26/118, 20.8%), and regression models for count data (Poisson + negative binomial 18/118, 15.3%). These choices are understandable under registry constraints and consistent with pre-pandemic influenza VE practice.(7,305) However, in a pandemic setting characterised by time-varying vaccination, rapid variant turnover, and shifting testing regimes, credible VE estimation required aligning time zero by calendar date/variant period so vaccinated and comparators enter risk simultaneously; using consistent risk windows (e.g., start ≥14 days post-dose) and censoring rules (e.g. booster receipt, infection, death); and harmonising testing opportunity (restrict or adjust for testing frequency).(23,306) Target-trial emulations provide this alignment, but were seldom used despite mature guidance.(24) Comparability was further limited by operational heterogeneity: definitions of ‘fully vaccinated’ (e.g., ≥ 7 vs ≥ 14 days post-dose), exclusion rules (e.g. prior infection, early post-dose person-time, immunocompromised individuals), and measures of association (odds ratios, relative risk and hazards ratios).

Adjustment and validation strategies were also notably uneven. Most studies adjusted for demographics, comorbidities, calendar time, and geography, but fewer incorporated variables closer to the principal threats in VE settings—testing behaviour and prior infection (selection into testing), exposure opportunity (household/occupation), and health-care utilisation. Several TND analyses additionally adjusted for health-care– seeking/utilisation; yet because TND already conditions on testing, further control for care-seeking can open collider pathways and bias VE estimates.(7,21) When comparability was enforced, exact matching (72/103, 69.9%) was favoured over weighting. Outside population-scale registries with dense covariate support, high-dimensional exact or strict matching, especially when it includes calendar date, can exclude large fractions of comparators when overlap is limited, thereby eroding power;(307) on the other hand, omitting calendar date risks residual confounding from differences in force of infection and variant mix.(308) Marginal weighting (stabilised IPTW or overlap weighting) preserves information and targets effects in regions of common support,(309) but was rarely used. Given that 155/253 (61.2%) of studies reported no matching/weighting at all, explicit weight diagnostics and positivity checks are needed. Additionally, about two-fifths of studies reported no sensitivity analyses overall, and negative-control outcomes/exposures or quantitative bias methods (e.g., E-values) were infrequently applied, despite accessible frameworks that make such checks feasible and interpretable for decision-makers,(26,310) which could reflect binding data constraints (limited capture of prior infection/testing, limited data on negative-exposure or outcome controls, incomplete linkages, sparse variant attribution).(308) Consequently, the limited use of validation checks leaves a non-negligible risk that some reported VE estimates are biased, either through residual confounding, study design or analytical strategy, which warrants cautious interpretation and privileging estimates with explicit identifying assumptions and documented robustness analyses.

Endpoints and comparators often aligned with programme priorities and objectives. As Omicron and its sublineages became predominant in 2023-24, baseline risks of severe outcomes declined;(138) together with smaller cohorts and shorter follow-up, this reduced power for rare endpoints (e.g. hospitalisation, ICU, death) and shifted attention toward infection or symptomatic disease, often as composites. As PCR testing and sequencing reduced, variant attribution increasingly relied on calendar-period proxies for dominant variant circulation, rather than sequence confirmation, limiting variant-specific severe-outcome analyses.(20,311,312) Early in the rollout, registry-rich settings such as Israel and England produced near-real-time estimates by linking vaccination, PCR, admissions, deaths, and sequencing; as emergency pipelines and legal authorities were withdrawn, this model became harder to sustain.(97,313) Comparator contrasts evolved with programme phases—initially one-dose or primary-series vs unvaccinated, later first-booster comparisons—while head-to-head platforms, heterologous schedules, bivalent-versus-monovalent boosters, and natural-immunity comparators remained uncommon. Platform focus also reflected supply and logistics: mRNA products were most evaluated in high-income settings, whereas inactivated vaccines were assessed more often in upper-middle-income settings, consistent with COVAX deliveries and uneven ultra-cold-chain expansion.(314,315) Future pandemic preparedness would benefit from more equitable VE evaluation capacity across income settings and continued assessment of updated vaccine formulations in endemic contexts.

This review has a few process limitations. This review was limited to peer-reviewed, English-language articles in PubMed/Ovid, which may bias toward high-income settings; however, only one additional non-English study (in Spanish) was identified and would have been excluded per eligibility criteria.(316) Meanwhile, focusing on routine administrative/registry data and excluding specialised cohorts was intended to keep data-generating processes comparable across jurisdictions, but may narrow generalisability to COVID-19 VE estimation methods in specialised populations. We did not preregister a synthesis protocol or apply ROBINS-I/GRADE, as the aim was to map methods rather than pool effects; however, data were extracted in duplicate with adjudication, study-level validity checks were catalogued across domains, and the extraction template, codebook, datasets, and analysis code are available to support replication.

Overall, the field delivered timely answers by leveraging available infrastructures, but with heterogeneity in design and robustness checks as the pandemic matured. A pragmatic way forward is not to discard familiar designs but to upgrade the workflow around them: **(i)** invest in routine, privacy-preserving linkage across vaccination, testing, encounters, vital records, and sequencing, especially in under-resourced settings; **(ii)** support VE research in underrepresented regions (e.g. LMICs) through capacity building and resource sharing to ensure findings are representative of global populations and healthcare contexts; **(iii)** pre-specify target-trial emulations where feasible and estimate marginal effects with inverse-probability or overlap weighting (or other doubly robust procedures); and **(iv)** make sensitivity and validity checks routine: at a minimum, confirm overlap (that vaccinated and unvaccinated groups exist across key strata), inspect depletion-of-susceptibles—particularly salient in later phases of the pandemic—by reporting VE stratified by prior-infection status (to distinguish true waning from faster infection among the unvaccinated), report E-values to appraise unmeasured confounding, and run negative-outcome control tests when data permits. Implemented together, these steps respond to the specific deficiencies documented here within the real data constraints, and could make observational VE estimates more transportable and decision-grade in future pandemics.

## Data Availability

Full-text extraction, supplementary codebook and related code used will be available on GitHub at first author's repository.

## Declarations

### Contribution statement

JYC and JTL conceived the study and developed the methodology. JYC, ZJG, RL, and DZYL performed screening and data extraction. All authors (JYC, ZJG, RL, DZYL, LEW, DCL, KBT, JTL) critically reviewed and edited the manuscript. JYC, LEW, and KBT contributed to literature search and writing of the manuscript. LEW, DCL, KBT, and JTL provided supervision. All authors had full access to the data and approved the final manuscript and the decision to submit.

### Data sharing

The template data-collection form and codebook are provided in the Supplementary Data S2. The study-level extraction dataset and the analysis-ready dataset used to generate all tables/figures are provided as Supplementary Data S3 (Excel). Analytic code (R scripts) will be publicly available on GitHub (repository link to be added).

